# COVID-19 management in social care in England: a systematic review of changing policies and newspaper reported staff perspectives

**DOI:** 10.1101/2021.11.17.21266410

**Authors:** Lavinia Bertini, Leanne Bogen-Johnston, Jo Middleton, Wendy Wood, Shanu Sadhwani, Julien Forder, Daniel Roland, Rebecca Sharp, John Drury, Jackie A Cassell

**Affiliations:** Department of Primary Care and Public Health, Brighton and Sussex Medical School, Falmer, UK; Evolution, behaviour and environment, School of Life Sciences, University of Sussex, Falmer, UK; Personal Social Services Research Unit, University of Kent, Canterbury, UK; School of Health Sciences, University of Brighton, UK; Kent Surrey Sussex Academic Health Science Network, Worthing, West Sussex, UK; School of Psychology, University of Sussex, Falmer, UK

## Abstract

Adult social care has been a major focus of public attention and infection control guidance during the COVID-19 pandemic, with a high mortality both for carers and those receiving care. To protect themselves and others from infection, staff in residential and domiciliary care settings had to quickly adapt to infection control measures that heavily impacted on their working and every-day life, whilst navigating new responsibilities, uncertainties and anxieties. We sought to explore the production and reception of guidance and look at ways these can be adapted to improve the working life of care staff in domiciliary and residential care whilst reducing the risk of SARS-CoV-2 transmission amid this pandemic and of future emerging infections.

We conducted two complementary and integrated systematic reviews of published documents in the pre-vaccination era: (1) National guidance for social care (conducted between 29 July to 28 October 2020), and (2) Newspaper coverage of infection control issues in social care (conducted between 27^th^ July to 10^th^ September 2020).

Three higher order common themes emerged in the integrated systematic review of guidance documents and newspaper articles: a) *Testing*, b) *Personal Protective Equipment*, c) *Employment*. The reviews revealed a sharp disjunction between the content of infection control guidance and its usability and applicability in social care settings. We suggest that infection control guidance needs to be better adapted to social care settings and informed by the sector. The practicalities of care work and care settings need to be at the core of the process for guidance to be relevant and effective. Modes and timings of communications also need to be optimised.

## 1. INTRODUCTION

Social care has been a major focus of activity and public attention during the COVID-19 pandemic. COVID-19 has presented a major challenge to adult social care settings, with a high proportion of all deaths among care home residents, and social care staff experiencing high death rates compared to other populations. Staff and people receiving care had to rapidly respond and adapt to new procedures, routines and guidelines, with new fears and responsibilities relating to infection and protection of infection for both care workers and those in receipt of care. In the UK, official figures on outbreaks and mortality in care homes became available from the end of March and early April. However people working in and representing the sector had voiced concerns in the media since the very early days of the pandemic (Hodgson et al., 2020).

In the UK, home care and care homes are part of the adult social care, which refers to a wide range of services and activities for adults (people aged 18+) to help people who are older and/or live with disabilities and physical or mental illness live independently, well and safely (The King’s Fund, 2019). In this paper, we focus on regulated social care for older people (65+) in care homes and domiciliary care, also referred to as home care (i.e., care in the person’s home).

On 9 March 2020, 33 care homes firstly reported COVID-19 outbreaks to Public Health England (PHE)^1^ (Hodgson et al., 2020) whilst the first death of a care home resident due to COVID-19 was registered on 17 March 2020 (Office for National Statistics, 2021). By 14 April, COVID-19 outbreaks had been confirmed in more than 2000 care homes - a third of all care homes for older people in England (Chan et al., 2021). Between 2 March 2020 and 12 June 2020, in England a total of 19,394 COVID-19 related deaths in care homes were reported, accounting for 29% of all deaths of care home residents (Office for National Statitstics, 3 July 2020). Care home deaths were 2.3 times greater than expected levels between 20 March and 7 May 2020 in comparison to the same period in previous years (Public Health England, 2020). From March to June 2020, 15.5% of deaths out of all hospital deaths were among care home residents (Office for National Statistics, 2020c). During this period, COVID-19 was the leading cause of death among male care home residents while it was the second leading cause among female residents (Office for National Statistics, 2020c). Still, these could be an underestimate of cases due to limited test availability during the first wave (Office for National Statistics, 2020c).

The Covid-19 experience of clients and staff in domiciliary care is harder to understand. While there was a rise in CQC notifications of death, these relate only to deaths during the delivery or as a result of regulated care(Office for National Statistics, 2020b).

Social care workers have also been disproportionately impacted by COVID-19. Between 21 March and 8 May 2020, 760 deaths occurred among care home staff in the 20 - 60 year age group, which was 346 deaths more than the same period in 2014 –2018 (Public Health England, 2020; Skills for Care, 2020) and twice the average during the same period in 2014 – 2019 (Office for National Statistics, 2020c). Moreover, the challenges of performing their duty of care whilst fearing cross-infection (Nyashanu et al., 2020)have brought an increase in signs of exhaustion, burnout, and staff shortages (Glynn et al., 2020).

In England, local authorities are responsible for commissioning social care, which is provided in the largest part by independent and autonomous businesses (National Audit Office, 2021). These providers are regulated, monitored and inspected by an independent regulator, the Care Quality Commission (CQC), which publishes ratings against a set of fundamental codes and standards of care (Cousins et al., 2021). The Department of Health and Social Care is responsible for setting national policy for adult social care. As such, domiciliary and residential care providers conform to advice from central and local governments whilst, economically, being run and managed as private businesses.

A key component of domiciliary and residential care is personal care, defined by the CQC as “supporting people in their homes (or where they’re living at the time) with things like washing, bathing or cleaning themselves, getting dressed or going to the toilet” (Care Quality Commission, 2012). These are often referred to as the activities of daily living (ADL) and involve close physical proximity between client and carer(s). Infection control in these social care settings for the elderly is paramount and complex due to a variety of interconnected factors, the most relevant being the high prevalence of comorbidity among those receiving care (Marshall et al., 2021; Tulloch et al., 2021) the type of care given and the physical proximity between cares and residents in care homes. Moreover, there is a high prevalence of cognitive impairment, including dementia, in the older population receiving residential or domiciliary care, with 60% of people receiving care at home and 69% of those in care homes living with dementia (Prince et al., 2014). As of June 2020, nearly half of care home deaths involving COVID-19 presented a diagnosis of dementia or Alzheimer’s disease (Cousins et al., 2021). Caring for people with dementia or Alzheimer’s disease posits important limitations and ethical considerations when implementing infection control measures like social distancing, shielding and wearing masks (Cousins et al., 2021; Nyashanu et al., 2020) as isolation and changes to routine and communication can be very distressing, with negative impact on their health and wellbeing (Cousins et al., 2021).

Despite its visible efforts and prompt response (Marshall et al., 2021) social care faced important delays in being able to implement recommended control measures including sparse testing and contact tracing, shortage of PPE, unclear protocols for discharging patients from hospitals into care homes (Cousins et al., 2021; Daly, 2020; Hollinghurst et al., 2021; Nyashanu et al., 2020). Moreover, although several governmental agencies and professional bodies have drafted guidelines specifically for care homes and domiciliary care during the COVID-19 pandemic (Sinclair et al., 2020; Wang et al., 2020) these have varied in quality and practicality.

Recommended COVID-19 related prevention measures included social distancing, wearing masks, improved sanitising behaviours, shielding and self-isolation. These can prove difficult to implement in domiciliary and residential care settings where carers and those receiving care are in close proximity. Moreover, workers in domiciliary care are likely to visit multiple clients a day, and clients may receive care from multiple care workers in diverse settings where ease of infection control practices varies enormously. Therefore, the domiciliary care sector may be unable to use all safety procedures that the care home sector has relied upon.

In this study, we aimed to explore the production of guidance, its reception and experience of using it in social care. We focused on the first wave of COVID-19 infections in the pre-vaccination phase of the pandemic to explore the challenges, barriers and enablers to the implementation of guidance in the care sector. This work forms part of a wider programme of work on Covid-19 in social care, including qualitative research and mathematical modelling.

## 2. METHODS

We conducted two complementary and integrated systematic reviews of documents in the pre-vaccination era: (1) National guidance for adult social care, and (2) Newspaper coverage of infection control issues in adult social care. This enabled us to explore the relationship between reported care worker experience and the guidance, as it was produced and communicated over time, during a period when methods and approvals for observational research required radical adaptation and faced significant delay.

Two systematic reviews were conducted by two researchers independently using PRISMA principles. One of guidance documents issued by policymakers and one of newspaper coverage of social care perspectives on managing COVID-19 transmission and risk of infection. Both reviews focused on the first wave of COVID-19 infections in the UK and addressed a common question: “what interventions and guidance will improve the working life of care staff in domiciliary and residential care whilst reducing the risk of SARS-CoV-2 transmission?”.

### 2.1 Guidance review

#### Search strategy

A systematic literature search for guidance documents was conducted within the electronic Government Digital Service, gov.uk, a United Kingdom public sector information website. The search was conducted on the 29 July 2020. Subsequent searches took place up until the 28 October 2020. Key terms “working guidance social domiciliary residential care staff SARS-COVID” were entered into the website search bar. Filters (Topic: Health and social care; Sub-topic: Social care; Content type: Guidance and regulation) were applied. Documents were included if they met the following criteria:

- Published 1 February to 28 October 2020
- Authored by the Department of Health and Social Care
- Referred to domiciliary care workers or/and residential care workers
- Offered guidance on improving working life of care staff during SARS-COVID-19 and/or reducing risk of SARS-COVID-19 transmission amongst care workers

Documents were excluded on the following criteria:

- Documents were not published between 1 February and 28 October 2020
- Documents were not authored by the Department of Health and Social Care
- Documents did not refer to SARS-COVID-19
- Documents did not refer to domiciliary care workers or/and residential care workers
- Documents did not include guidance on improving working life of care staff during SARS-COVID-19 and/or did not included guidance on reducing risk of SARS-COVID-19 transmission amongst care workers.

Documents were excluded on title. The sample was refined further after a cursory read. Remaining documents were read in-depth until the final sample met inclusion criteria. During reading, documents that were cited by, but had not been included in, the data set were explored. These were included if they were found to meet inclusion criteria. The URL of each selected document was saved into Wayback Machine (https://archive.org/web/), a digital archive of the World Wide Web, allowing for the search of previous and updated versions of each document. Versions were searched via the Wayback Machine ‘Changes’ tool which identifies, and displays, “changes in the content of archives of URLs” (Wayback Machine, 15 November 2020). URLs were compared (two at a time) side-by-side. Changes were highlighted in blue (information added) and yellow (information removed). Each updated version was included in the data set.

### 2.2 Newspaper articles review

#### Search strategy

A systematic review of newspaper articles was conducted from July to September 2020, with the first search taking place on 27^th^ of July and the last on 10^th^ of September. The review focused on newspaper articles that reported, either directly or indirectly, views and experiences of social care managers, care home staff and domiciliary care staff on managing SARS-CoV-2 in their working environments. Articles were included on the following criteria:

- published between 1st of July to 31st of August 2020
- reported, quoted or included views from social care workers and/or managers and/or sector representatives
- written in English
- published either nationally or locally in Kent, Sussex, and Surrey

Articles were not included for review if:

- they were not published between 1st of July to 31st of August 2020
- they did not report, quote or include views from social care workers and/or managers and/or sector representatives
- they were not written in English
- they analysed or summarised government guidelines, briefings or reports but did not relate direct quotes or views from social care staff and representatives.

The systematic review was carried out using the online news research database tool Nexis®, with the search terms: (“domiciliary care” OR “care homes” OR “social care”) AND (“covid-19” OR “COVID” OR “coronavirus”) AND (“care home managers” OR “care workers” OR “care staff”). Searches were conducted in English. The selection process is shown in Figure 1.

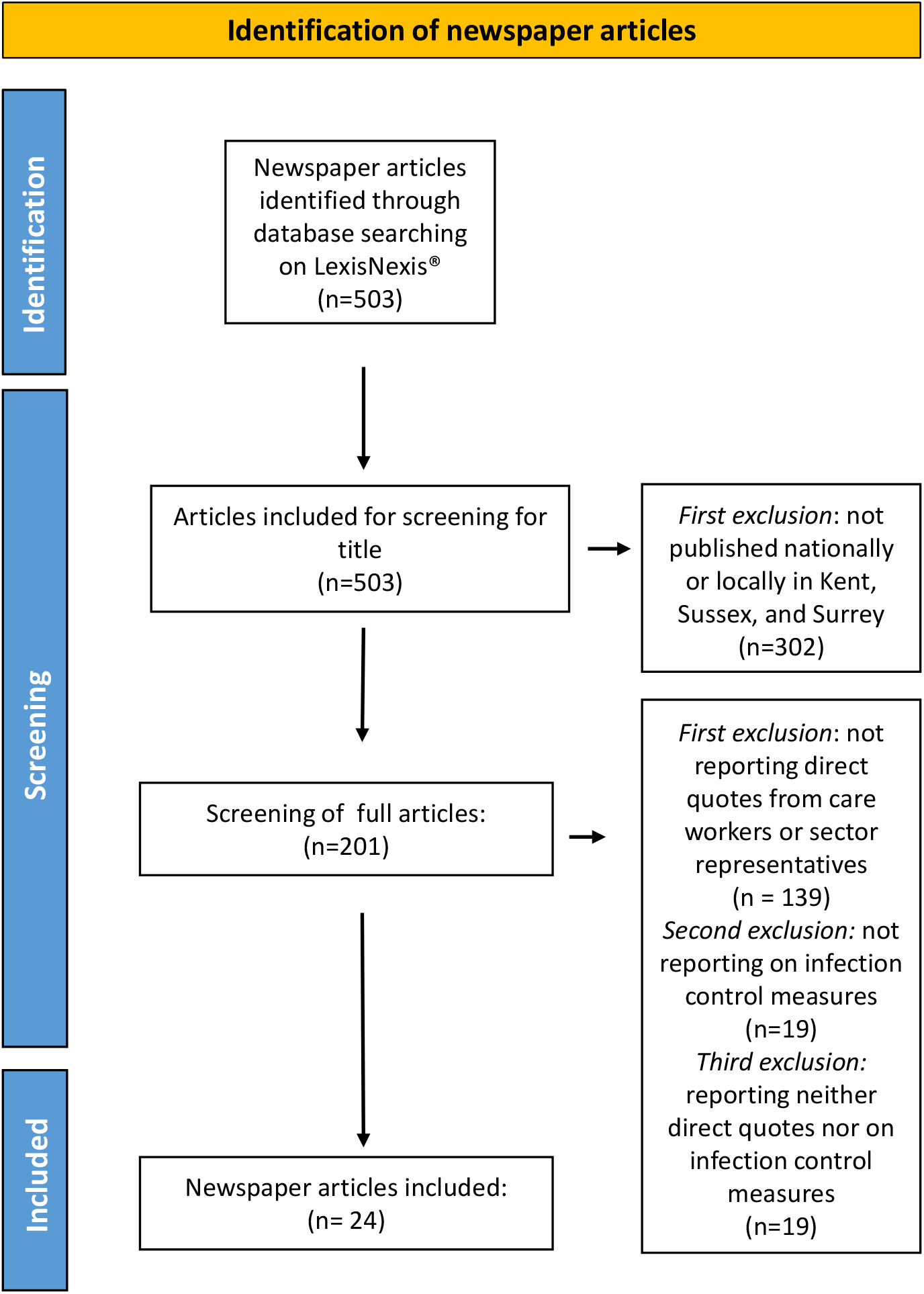

### 2.3 Data Analysis

Data from both reviews were analysed through Framework Analysis (Gale et al., 2013; Ritchie & Spencer, 2002). Each document was read twice for familiarisation and relevant sections coded. To manage and organise data, preliminary codes and sections of data were entered into a Microsoft Excel 2016 spreadsheet. A preliminary framework was developed from the initial codes. Data within each cell were summarised. Codes were then grouped into themes and higher order themes. Temporal differences within the data from the systematic review of guidance were compared to track updates and changes in official guidelines. The framework was refined and compared against the research question for meaning and significance. Analysis was inductive and exploratory, allowing novel themes and issues to emerge from the data.

## 3. RESULTS

We identified three higher order common themes in the systematic review of guidance documents and newspaper articles: a) *Testing*, b) *Personal Protective Equipment*, c) *Employment*.

### 3.1 Guidance review

The literature search produced 262 result. After inclusion and exclusion criteria were applied a total of 15 government documents (excluding updated versions) were included (see Table 1).

**Table 1.** List of government documents included for review

| Code | Author | Policy Name | Versions/Updates |
| --- | --- | --- | --- |
| GD1 | Department of Health and Social Care | Covid-19 Information governance advice for the social care sector | **1 April 20**<br>4 May 20<br>6 June 20<br>4 July 20<br>2 Oct 20 |
| GD2 | Department of Health and Social Care | Admission and Care of Residents in a Care Home during Covid-19 | **2 April 20**<br>20 April 20<br>19 May 20<br>19 June 20<br>31 July 20<br>14 August 20<br>27 Aug 20<br>2 Sept 20 |
| GD3 | Department of Health and Social Care | COVID-19: management of staff and exposed patients or residents in health and social care settings | **4 April 20**<br>17 April 20<br>16 May 20<br>14 June 20<br>31 July 20<br>18 Aug 20<br>28 Sept 20 |
| GD4 | Department of Health and Social Care | Coronavirus (COVID-19): adult social care action plan | **16 April 20** |
| GD5 | Department of Health and Social Care | Personal protective equipment (PPE) – resource for care workers delivering homecare (domiciliary care) during sustained COVID-19 transmission in the UK | **27 April 20**<br>30 April 20<br>1 June 20<br>15 June 20<br>20 July 20<br>28 Aug 20<br>29 Sept 20<br>7 Oct 20 |
| GD6 | Department of Health and Social Care | Health and wellbeing of the adult social care workforce | **11 May 20** |
| GD7 | Department of Health and Social Care | Government to offer antibody tests to health and social care staff and patients in England | **22 May 20**<br>17 Sept 20 |
| GD8 | Department of Health and Social Care | Coronavirus (COVID-19): provision of home care | **22 May 20**<br>24 June 20<br>7 Aug 20<br>2 Sept 20 |
| GD9 | Department of Health and Social Care | Dedicated app for social care workers launched | **6 May 20** |
| GD10 | Department of Health and Social Care | Coronavirus (COVID-19): care home support package | **19 May 20**<br>22 May 20<br>9 July 20 |
| GD11 | Department of Health and Social Care | COVID-19: Adult Social Care Risk Reduction Framework: Assessing and reducing the risk to your workforce | **10 July 20** |
| GD12 | Department of Health and Social Care | Update on policies for visiting arrangements in care homes | **22 July 20**<br>21 Sept 20<br>15 Oct 20 |
| GD13 | Department of Health and Social Care | COVID-19 Guidance on redeploying workers and involving volunteers | 4 Aug 20 |
| GD14 | Department of Health and Social Care | Overview of adult social care guidance on coronavirus (COVID-19) | 25 Aug 20<br>26 Aug 20<br>11 Sept 20 |

#### 3.1.1 Reducing Transmission Between Spaces

##### Care Workers

To constrain the asymptomatic transmission of infection, The Department of Health and Social Care (DHSC) looked to limit workforce movement between care homes, requiring providers to ensure that all care homes and agency staff work at a single location. It was recommended that staff movement (if agreed with the staff member) within the care home should also be restricted by patient group (COVID-positive or COVID-negative) (GD10, 19 May 2020). This was reiterated in the 2020 to 2021 winter plan (GD15, 18 Sept 2020). Similarly, to reduce numbers of workers entering a home, domiciliary care providers were recommended to create a multi-agency approach and allocate staff to ‘care groups’ (GD8, 22 May 2020).

Care home providers were asked to provide accommodation, either on site or with local hotels, for staff who wished to minimise social interaction (GD2, 19 June 2020). Guidance also suggested that staff should be encouraged and supported to avoid the use of public transport (GD2, 19 June 2020).

Implementing such restrictions, would add financial burden for care providers e.g. recruitment of additional staff (GD10, 19 May, 2020). To meet costs, adult social care providers were advised to access funding from an additional £600 million, Infection Control Fund (announced on 14 May 2020) (GD8, 22 May 2020).

##### Residents and Clients

On 16 April 2020, the government introduced testing for all residents prior to care home (re)admission. Those awaiting test results would still be admitted but, to safeguard staff and residents, isolated if the care home were able to accommodate this (GD4, 16 April 2020). In September 2020, updated guidance stated that anyone with a COVID-positive test, due to be discharged from hospital to a care home, would first be discharged and cared for at a designated infection-controlled accommodation for the required isolation period. Patients with an outstanding COVID-19 test or without having been tested within the past 48 hours would not be discharged to a care home (GD15, 18 September 2020).

In respect of those in receipt of domiciliary care, regardless of COVID-19 status, individuals discharged from hospital could be cared for at home by care workers, providing PPE was adhered to (GD8, 22 May 2020).

#### 3.1.2 Protecting from Infection

##### Contact Between Workers

The Government issued guidance on protecting care workers against contracting COVID-19. One approach was to decrease contact between domiciliary workers. Interaction between staff (e.g. team meetings and handovers) should be conducted remotely and “teams and individuals should have remote access to regular supervision” (GD8, 22 May 2020).

##### Testing

The government also looked to increase staff testing and outlined that:

All symptomatic social care workers, including care home staff, have been able to access a test since 8 April and PHE have been providing testing to support outbreak control in care homes since the start of the outbreak (Admission and Care of Residents in a Care Home during COVID-19, 19 June 2020).

##### Personal Protective Equipment (PPE)

While guidance (GD2, 2 April 2020) emphasised the importance of PPE, global demand in April 2020 resulted in shortages with many care providers unable to procure PPE through usual channels. Despite the government response of a National Supply Disruption Response system in April (GD4, 16 April, 2020) and an emergency PPE portal system in June (GD14, 25 August 2020), it was not until September 2020 that reference to acute PPE shortage guidance was removed from government policy (GD14, 11 September 2020).

##### Exposure

In cases where a worker had been exposed to COVID-19, guidance stated that:

Health care workers who come into contact with a COVID-19 patient or a patient suspected of having COVID-19 while not wearing personal protective equipment (PPE) can remain at work (GD3, 4 April, 2020).

On the June 14th, policy was updated whereby a care worker exposed to COVID-19 whilst providing personal care to a resident or service user should undertake a risk assessment in conjunction with local infection prevention and control policy. Workers should not attend work if a significant breach had occurred. This also applied for staff who had contact with a co-worker confirmed as a COVID19 case (GD3, 14 June 2020).

#### 3.1.3 Staff Wellbeing

Guidance issued by the government also mentioned staff wellbeing: “Providers should ensure that there is a high level of support and a focus on staff health and wellbeing” (GD8, 22 May 2020).

##### Financial Wellbeing

In March 2020, £1.6 billion of additional funding was granted to local authorities. Part of this funding was to financially support care workers (a quarter of whom were on zero-hour contracts) unable to work for short periods of time due to ill health or self-isolation. Care workers unable to work for longer periods (high-risk groups) were to be furloughed, receiving 80% of normal income. Increases in the basic rate of Working Tax Credit and the standard allowance in Universal Credit were designed to help those (in receipt of benefits) working extra shifts (GD4, 16 April 2020).

Regarding redeployment, employers were advised to obtain agreement by the worker (GD13, 4 August 2020), to ensure that contractual arrangements were suitable and in the “best interests of the worker” and develop a plan for managing seconded workers. While current pay, terms and conditions were to remain the same, receiving employers could (if they wished) pay above a worker’s usual pay (GD13, 4 August 2020).

##### Mental and Physical Health Wellbeing

Digital resources, such as a free text messaging service, were developed to aid care worker wellbeing. In April 2020 a dedicated website, and a helpline to support wellbeing, were in development (GD4, 16 April 2020). The CARE Workforce website and app were introduced in May 2020. Advice on how employers could help staff wellbeing was also published (GD8, 22 May 2020). Adult social care guidance for winter (GD15, 18 September 2020) advised that workplace risk assessments should be undertaken by employers with staff. ‘A risk reduction framework for adult social care’ offered employers guidance on discussing risks with potentially vulnerable staff.

### 3.2 Newspaper articles review

The search gave 503 results of which 24 were included for analysis after screening (see Figure 1). The articles included are listed in Table 2. The following themes emerged from the analysis.

**Table 2.** List of newspaper articles included for review

| Code | Title | Author | Newspaper | Date |
| --- | --- | --- | --- | --- |
| NPA1 | *Coronavirus: Weekly testing for care home staff to start on Monday; Change in strategy follows advice from Sage and new study showing higher prevalence of virus in care settings.* | Shaun Lintern | The Independent (Web Publication) | 03/07/20 |
| NPA2 | *Staff in care homes to have coronavirus test every week.* | Shaun Lintern | The Independent | 03/07/20 |
| NPA3 | *Boris Johnson refuses to apologise for blaming care homes for coronavirus death toll; Claim that care homes did not follow correct procedures branded 'insulting' to staff and operators.* | Andrew Woodcock | The Independent | 07/07/20 |
| NPA4 | *Piers Morgan calls Boris Johnson 'disgusting' for blaming care homes for coronavirus deaths; Government attempting to 'shift' the narrative, says Morgan.* | Matt Mathers | The Independent | 07/07/20 |
| NPA5 | *'Travesty of leadership': Charity boss hits out at 'cowardly' Boris Johnson after PM blames care homes for coronavirus.* | Ashley Cowburn | The Independent (Web Publication) | 07/07/20 |
| NPA6 | *325 sent to care homes without C-19 test.* | Jez Hemming | The Western Mail | 08/07/20 |
| NPA7 | *Downing St refuses to comments 'real slap in apologise after PM's face for care workers'.* | Harriet Line | The Western Mail | 08/07/20 |
| NPA8 | *Slap in the face from PM as 405 die in care homes.* | Nicole Jordan | Medway Messenger | 09/07/20 |
| NPA9 | *Coronavirus: Treat relatives of care home residents with dementia as key workers, charities say; Department of Health and Social Care says it will release details on allowing care home visits 'shortly'.* | Emily Goddard | The Independent (Web Publication) | 10/07/20 |
| NPA10 | *Coronavirus: UK frontline worker death rate second highest among 79 countries, report shows; At least 540 staff have died in England and Wales, Amnesty International says.* | Emma Bowden | The Independent | 13/07/20 |
| NPA11 | *'Emotionally, I have on many occasions gone home feeling completely destroyed...'; In an extensive survey, frontline healthcare workers reflect on Wales' handling of the coronavirus pandemic so far.* | Mark Smith | The Western Mail | 16/07/20 |
| NPA12 | *Ministers accused of 'sidestepping' issue of low pay for social care workers after nearly 1m other key staff get pay rises; Liberal Democrats say omission is 'simply not acceptable'.* | Ashley Cowburn | The Independent | 21/07/20 |
| NPA13 | *NO HELP FOR NHS HEROES; Frontline staff suffer pay snub.* | Jack Andrews | Daily Star | 22/07/20 |
| NPA14 | *More than 12,000 people demand care worker be allowed to stay in country.* | Jody Doherty-Cove | The Argus | 30/07/20 |
| NPA15 | *Fury after care homes go without promised testing - MP speaks to PM directly.* | Jody Doherty-Cove | The Argus | 31/07/20 |
| NPA16 | *Care worker lost husband to Covid-19 - and now needs thousands of pounds to guarantee family can stay in UK; 'Free and automatic indefinite leave to remain would give me space to grieve,' she says.* | Zoe Tidman | The Independent | 02/08/20 |
| NPA17 | *Coronavirus: Ministers accused of 'negligence' towards care home residents and staff; Rollout of regular Covid-19 testing delayed because of problems with supplies.* | Andrew Woodcock | The Independent | 02/08/20 |
| NPA18 | *Bereaved care worker has to pay thousands of pounds to guarantee UK residence.* | Zoe Tidman | The Independent (Daily Edition) | 03/08/20 |
| NPA19 | *Now the clapping has stopped, let's be ready to fight a pandemic of mental ill health among NHS staff; It's no surprise that the mental health of NHS staff is under increasing strain. If we are to face a second wave of coronavirus, then these key workers must be given better support, says Jennifer Hawkins.* | Jennifer Hawkins | The Independent | 12/08/20 |
| NPA20 | *We need a government minister dedicated to supporting our ageing population; If we as a society can't fix how we look after our vulnerable when their lives are threatened by a pandemic, I cannot imagine when we ever will. Something needs to change, writes Sanjeev Kanoria.* | Sanjeev Kanoria | The Independent | 17/08/20 |
| NPA21 | *Ageism means we need a minister for older people.* | Sanjeev Kanoria | The Independent (Daily Edition) | 18/08/20 |
| NPA21 | *Supported living test 'disgrace'.* | N/A | The Western Mail | 20/08/20 |
| NPA23 | *Coronavirus: Study reveals alarming impact of Covid on care home sector; One nurse tells study: 'We were under constant pressure to admit people who were Covid positive.* | Shaun Lintern | The Independent | 22/08/20 |
| NPA24 | *Coronavirus: 'Confusing' advice from Public Health England put patients at risk, watchdog says; One vulnerable patient died after care workers visiting their home were told they did not need to wear PPE.* | Shaun Lintern | The Independent | 26/08/20 |

#### 3.2.1 Testing

Issues and comments about testing in social care settings were the most reported (n=10) in the articles analysed and referred primarily to the introduction of weekly testing for care staff and monthly testing for residents in care homes. A couple of articles reported positive comments on new testing guidelines.. However, most articles report concerns from sector representatives lamenting that advice and implementation of testing arrived too late (end of May) and only after continuous pressure from the sector.

Two articles (NPA15 and NPA17) relay the direct experience of a national social care providers and local spokesman reporting delays in receiving test kits in July and beginning of August.

Two newspaper articles also reported about the need to test care staff more than once, as explained by Mark Adams Chief Executive of the Community Integrated Care charity: “once is absolutely useless because if you get tested and then get back on the bus and pick up the virus on the bus, within a week you’re potentially asymptomatic and infectious” (NPA4).

One article reports the words of an anonymous staff nurse commenting on the pressure on care homes to receive patients without testing from hospitals as well as the lack of clear guidance on returning to work for staff who tested positive: “I unfortunately had the virus and there was no advice from my occupational health department, except that I could go back to work on day eight if better” (NPA23).

#### 3.2.2 Employment

Issues related to employment represented the second higher theme (n=7). These were often mentioned when discussing wider issues that had a negative impact on the sector during COVID-19 but preceded the pandemic. Two main sub-themes were identified: *visa regulation and employment rights*.

##### Visa regulation

Issues around visa regulation and it impacts on care staff were reported by 4 articles (NPA14; NPA16; NPA18, NPA20). These were described through the stories of two care workers who lost their right to work and live in the UK whilst they were working in social care during the pandemic. Dr Sanjeev Kanoria owner of Advinia HealthCare, one of the largest private care providers in the UK, comments: “Care workers have also been excluded from a post-Brexit fast-track visa system for health workers” (NPA20).

##### Employment rights

The issue of the majority of care staff being on minimum wage and lack of access to basic rights, such as sick pay, was reported by three articles where sector representatives described this situation as “unacceptable” (Vic Rayner, executive director at the National Care Forum, in NPA12) and a “matter of national shame” (Mark Adams, Head of social care charity Community Integrated Care, in NPA13).

#### 3.2.3 Guidance

From the articles analysed, issues around guidance also emerge as a main concern. Six articles (n=6) report extensive comments on issues around guidance. The main issues identified are: a) the delay in providing care homes with guidance; b) guidelines being ever-changing and changes communicated late (often on a Friday night); c) guidance based on secondary care with little understanding of care services, hence ineffective or counter-productive.

National Care Forum’s executive director Vic Rayner, referring to guidance says that “government guidance has come to the sector in stops and starts - with organisations grappling with over 100 pieces of additional guidance in the same number of days, much of which was not accompanied by an understanding of the operational implications of operating care services” (NPA15). Jane Townson added that lack of clarity “led to inconsistent local interpretations being made by public health, local councils and community health organisations” and that “dissemination was woefully inadequate” (NPA15). She also suggests that including those who know social care in the drafting process could be a positive step to address some of these issues.

Delays on producing and receiving guidelines on visits to care homes was mentioned only in one of the selected articles, by Martin Green, chief executive of Care England who also supported the call to treat relatives of care home residents as key workers (NPA9).

#### 3.2.4 Personal Protective Equipment (PPE)

Insufficient PPE and lack of support in providing PPE is also reported in the article analysed, although to a lesser extent (n=5) than issues around testing and guidance. The articles that directly mention this issue tended to represent it as a well-known and verified fact and the comments reported were often lengthier and more detailed than those for higher themes. Both lack of PPE and quality of PPE were relayed as cause for concern. Jane Townson, chief executive of the United Kingdom Homecare Association, commented that: “The production of official guidance on personal protective equipment has been a shambles throughout the coronavirus pandemic.” (NPA24). An anonymous nurse in a study reported by The Independent (NPA24), said that “it took seven weeks just to get staff masks properly tested to ensure they were safe” and “We had no proper support until the end of May.” The Independent Care Group’s chairman Mike Padgham also highlighted that care staff had to deal with “late and conflicting advice and poor support in terms of personal protective equipment” (NPA7).

#### 3.2.5 Feelings of neglect and blame

Newspaper articles (n=4) also reported the reactions sparked by Prime Minister’s comment that the high numbers of deaths in care homes was due to care homes not following government’s advice and guidance. Sector representatives describe it as “unacceptable” (NPA4) and “a slap in the face” (NPA7). These reactions speak of a sector that feels to be wrongly blamed and overlooked in dealing with the pandemic. Chief executive of Care England Martin Green (NPA8) and the chief executive officer of Community Integrated Care, Mark Adams (NPA5), accompanied these reactions to comment on the lack of leadership and accountability from the government in the decision process.

#### 3.2.6 Sector reform

A smaller number of newspaper articles (n=3) reported on the need to reform the sector, often mentioned in connection to issues of pay and staff shortage (including visa/work regulation). The sector representatives cited in the articles (NPA7; NPA20; NPA21) lamented that reform of social care is long overdue and has been debated for over a decade. The Independent Care Group’s chairman Mike Padgham described it as “long-promised” and one of the main reasons that made the sector so vulnerable to Covid-19 (NPA7). Dr Sanjeev Kanoria owner of Advinia HealthCare, affirmed that “the [social care] system that entered the pandemic was underfunded, understaffed, undervalued and in need of fundamental reform” (NPA20).

## 4. DISCUSSION

The reviews revealed a sharp disjunction between the content if infection control guidance and its usability and applicability in social care settings. It is also important to note that this disjunction between guidance and its applicability is often produced by guidance not taking into account ongoing systematic and logistical barriers encountered by the social care sector, such as limited supply chains of PPE and testing kits (Daly, 2020; Marshall et al., 2021; Nyashanu et al., 2020), staff shortage and precarious employment (Daly, 2020; Marshall et al., 2021).

The disjunction between guidance and its applicability is evident around testing, which emerged as a higher order theme in both reviews. Testing has been deployed as a key measure both to reduce transmission across setting and prevent infection. However, findings show that newspaper articles reported providers’ difficulties in accessing testing kits for staff and residents even some months after testing in care homes was introduced in governmental guidance. The same issues around delays and access to testing kits for staff and clients are also evident in the analysis of policy guidance and documented by existing literature (Nyashanu et al., 2020). Despite a government announcement that all symptomatic care workers could obtain a test since 8 April^6^, due to limited capacity it was not until the 28 April that all care home workers were in practice entitled to a test (National Audit Office, 2020). However, the daily number of care home tests were not only capped at 30,000, but split between staff and residents (National Audit Office, 2020).

The testing and isolation of patients (re)admitted to care homes after being discharged from hospital represents another important area where guidance proved difficult to implement. Plans for the (re)admission of residents from hospitals to care homes were updated in September 2020, with the proposal of designated infection-controlled accommodation. However, establishing infection-controlled homes for discharged COVID-positive patients has proven problematic, with many councils failing to nominate an appropriate location by the required October 2020 deadline (Booth, 2020b). In addition, this raises concerns as to whether it is ethical to ask “staff to place themselves in the way of potentially contracting the virus”(Booth, 2020a).

The implementation of infection control measures on testing and isolation of residents and staff requires a larger workforce (Daly, 2020). For example, to limit the transmission of infection across spaces, guidance asks that staff work at a single location and that staff movement within the care home and domiciliary care is restricted by patient group. These measures require an increase in staffing ratios (Gordon et al., 2020) only possible if the sector could rely on high volumes of trained workforce and recruitment. Increase staffing can prove particularly challenging at a time where staff have to self-isolate if symptomatic or might be shielding. The need for a higher workforce to implement infection control measures is also at odds with longstanding and well-known issues around workforce shortages^2^, staff retention and low pay that the social care sector has been facing for over a decade (Marshall et al., 2021; National Audit Office, 2021). The government granted extra funding (March 2020 and May 2020) that could be in part use by local authorities and care providers to meet costs for extra staffing and support furlough of staff who couldn’t work (e.g., staff in high-risk group). An economic evaluation of the effectiveness of such extra funding is beyond the scope of this review. The review highlights the inadequacy of incentives and safeguards for undertaking social care work during a pandemic. For example, results from both reviews show that little was made to guarantee fairer wages, improved rights (e.g., sick pay), relaxed visa regulation or mental health support for care workers. At this end, it is important to note that 24% of adult social care workforce and 43% of domiciliary care workforce are on zero-hour contracts (Skills for Care, 2020), that do not guarantee a basic income or basic rights. Others have a reported an association between homes not providing staff with sick pay and COVID-19 infection in residents (Tulloch et al., 2021), suggesting that guidance failed to optimise the support and protection required by adult social care workers during the first wave of COVID-19 pandemic, before the dramatic impact of vaccination had emerged.

A final point that emerges from the reviews is the need to optimise the content and dissemination of guidance. The main issues reported by sector representatives are: high volume of guidelines; inadequate and ever-changing guidance; lack of effective and timely communication; guidance based on secondary care with little understanding of care services. The data visualised in Table 4 show the changes and updates made to infection control guidance between April and October 2020, supporting the sector’s concern for ever-changing guidance. Moreover, guidelines were deemed to be designed around NHS as a model and to show poor understanding of the organisational and operational specificities of residential and domiciliary care. Guidance was often released on a Friday night and at very short notice from its implementation, “with organisations grappling with over 100 pieces of additional guidance in the same number of days” (NPA15). It is likely that this translated into added work for organisations and managers to make ever-changing and overly long guidance accessible to staff and operational. It is also important to note that guidance focused mainly on care homes, suggesting that there was limited attention given to domiciliary care among policy makers, with important consequences for the safety of staff and those receiving care.

The communication problems identified here – delay, constantly changing guidance, and lack of understanding of needs in the care sector – echo some of those identified more generally across the pandemic (Independent SAGE, 2020). For example, the experience of receiving frequent and ever-changing guidance had the effect of undermining both public understanding of the rules and their trust in the authorities (Hill et al., 2020). Research on other public health emergencies suggests that where provision of information is seen as timely rather than delayed, this enhances the whole relationship between providers and receivers of information, as well as reducing anxiety (Carter et al., 2015). A possible solution to the problem of guidance not being calibrated to the needs of care sector lies perhaps in principles of co-production, in which the guidance is created together between ‘decision-makers’ and practitioners – something recommended in the UK context by SPI-B (2021) the behavioural science subgroup of SAGE, as an overall strategy for control of Covid-19.

### 4.1 Recommendations

We suggest that guidance on infection control in social care settings should be informed by the practical knowledge of those providing and receiving care, possibly through co-production (Bear et al., 2021; Durose et al., 2017). This would ensure that the practicalities of care work and care settings are adequately understood and incorporated so that guidance is relevant, effective and applicable. We further suggest that infection control guidance for social care must be centred on the understanding of care settings as places where residents and clients live and socialise (Stone et al., 2015), unlike hospitals.

A second recommendation concerns the importance of supporting the workforce. We suggest that understanding the implications of precarious employment in social care is fundamental for the production and implementation of infection control guidance. Precarious working conditions and low pay are likely to have an important impact on workers’ choices and possibilities to adopt COVID-19 measures, for example in relation to shielding, self-isolating, working for multiple care agencies, demanding safer working practices, etc. Any guidance that overlooks or dismisses these aspects run the risk of being ineffective and/or encouraging dynamics potentially harmful for workers and residents or clients. Measures and incentives aimed at supporting and safeguarding workers directly, such as pay rise, sick pay, relaxed visa regulations, should be considered integral part of infection control guidance. This is particularly relevant when guidance is produced for low-paying sectors with “increasing precarious working arrangements” (Hussein, 2017, p. 1817).

We suggest that it is also important to listen to the general (Hussein, 2017, p. 1817)sentiment of neglect voiced by the sector. We have already mentioned how social care settings felt neglected at the beginning of the pandemic, with government’s efforts on testing and PPE being primarily focused on the NHS. The inadequacy of guidance for care homes and home cares due little understanding of these settings as discussed earlier, is also imbued with feelings of neglect and being unfairly blamed. Comments around guidance, testing, and PPE as well as pay rise and bereavement have in common the fact that where the government had implemented positive initiatives, care workers were never included in the initial plans and only managed to get access to some of these after a sector and/or public outcry.

### 4.2 Limitations

Two main limitations of this study are the narrow geographical and temporal focus of the systematic review of newspapers articles, limited to local newspapers in Kent, Surrey and Sussex published between 1^st^ July and 31^st^ August 2020, alongside national coverage. This is due reasons of resource and funding focus, as these systematic reviews were developed as part of a broader project exploring the impact of COVID-19 infection control guidance on the working lives of domiciliary and residential care staff in Kent, Surrey and Sussex.

We are also aware that the issues analysed in this review and reported in newspapers are mediated through journalists and editorial boards. As such, the review does not report the direct experiences and opinions of sector representatives. A qualitative media analysis review of newspaper and/or media coverage of staff perspectives on COVID-19 infection control measures throughout the pandemic might add further insight to the findings presented in this research.

Moreover, it is important to note that the majority of the newspaper articles included in the review report comments from sector representatives and care providers whilst the direct voices of carers are less present. It would be fruitful to compare the themes identified in this review with those that might emerge from interviews and/or focus groups with domiciliary and residential care staff on their experience of implementing covid-19 infection control guidance.

## Data Availability

All data produced in the present study are available upon reasonable request to the authors

## Acknowledgements

We would like to thank our colleague Dr. Hasarali Fernando (Brighton and Sussex Medical School) for her input to identify relevant epidemiological data for this paper.

---

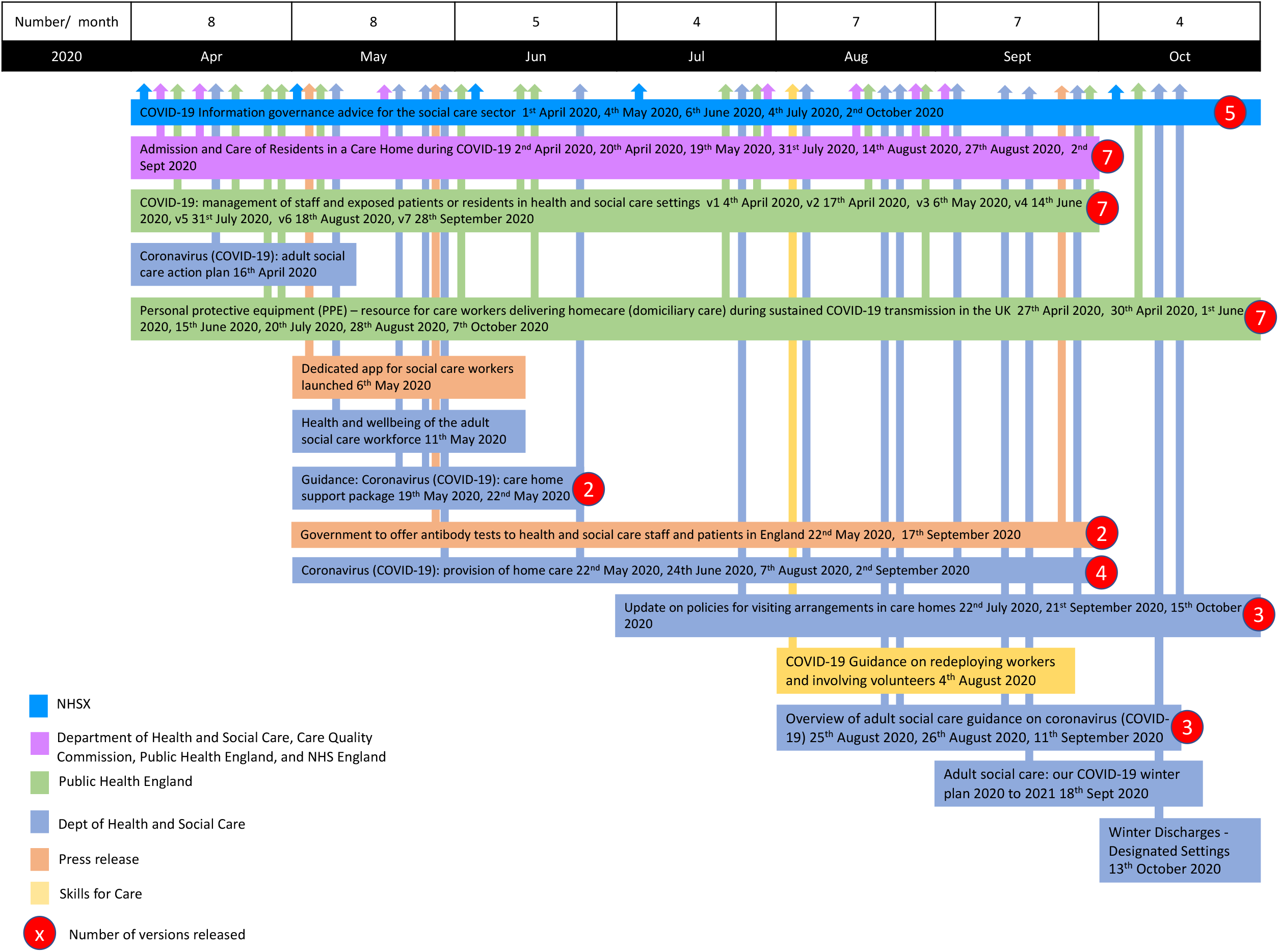

## Footnotes

1 Public Health England is responsible for monitoring outbreaks in care homes

2 It is estimated that in 2019/2020 there were approximately 112,000 vacancies at any one time, equivalent to 7.3% of the roles in adult social care (Skills for Care, 2020)

